# Filling the gaps: leveraging large language models for temporal harmonization of clinical text across multiple medical visits for clinical prediction

**DOI:** 10.1101/2024.05.06.24306959

**Authors:** Inyoung Choi, Qi Long, Emily Getzen

## Abstract

Electronic health records offer great promise for early disease detection, treatment evaluation, information discovery, and other important facets of precision health. Clinical notes, in particular, may contain nuanced information about a patient’s condition, treatment plans, and history that structured data may not capture. As a result, and with advancements in natural language processing, clinical notes have been increasingly used in supervised prediction models. To predict long-term outcomes such as chronic disease and mortality, it is often advantageous to leverage data occurring at multiple time points in a patient’s history. However, these data are often collected at irregular time intervals and varying frequencies, thus posing an analytical challenge. Here, we propose the use of large language models (LLMs) for robust temporal harmonization of clinical notes across multiple visits. We compare multiple state-of-the-art LLMs in their ability to generate useful information during time gaps, and evaluate performance in supervised deep learning models for clinical prediction.

## Introduction

In 2009, the United States introduced the Health Information Technology for Economic and Clinical Health (HITECH) Act with the aim of promoting the implementation of health information technology^1^. Since then, there has been a significant increase in the utilization of Electronic Health Records systems (EHR), which are digital versions of a patient’s medical chart. These data, which include information about a patient’s medical history, diagnoses, medications, laboratory test results, and treatment plans, hold great potential for application in various healthcare settings. EHR data have been employed for various purposes such as disease prediction, risk assessment, and assisting clinical decisions. Clinical notes in particular, which may contain important contextual information regarding family history, subtleties of symptoms and treatments, early warning signs, lifestyle, and socioeconomic factors, have been increasingly used for predictive modeling of disease.^2-15^

However, EHR presents several challenges as it contains data that is recorded at irregular intervals and with varying frequencies (Figure 1)^16^. This can happen for a variety of reasons– in an emergency department (ED) or intensive care setting (ICU), patients with more frequent visits might be sicker than their counterparts.^17-18^ In other settings such as primary care, patients from different groups may have differing levels of access to healthcare. For example, studies have found that underserved populations are more likely to visit multiple healthcare institutions to receive care, thus contributing to data fragmentation^19^. It has also been found that disproportionate missing data in patients within certain demographic groups deteriorates the prediction of disease, emphasizing the impact that missing data may have on underserved populations^20^.

**Figure 1:**
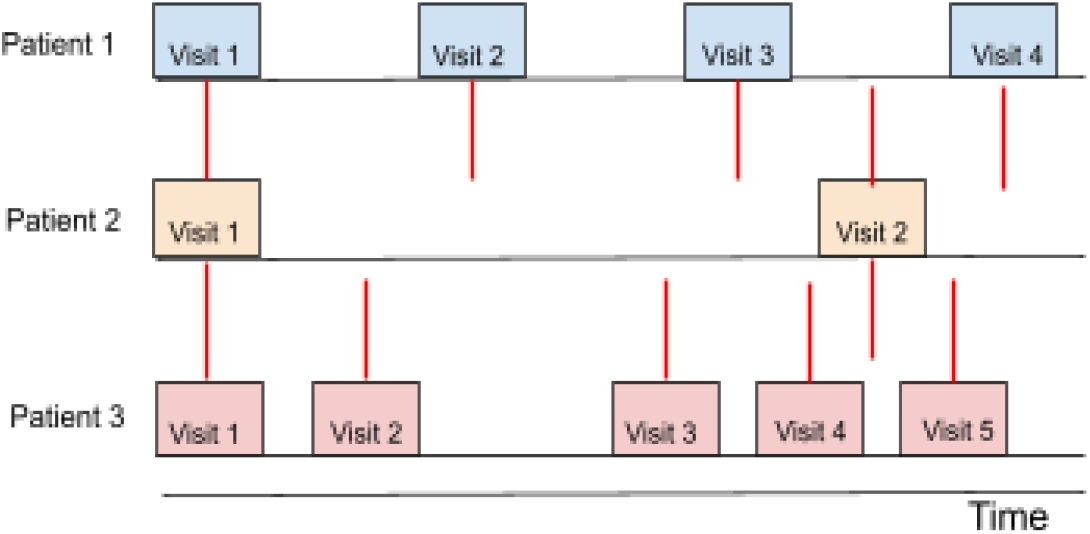
Medical visits occur at irregular time intervals and varying frequencies.

Deep learning models that can handle time series data such as Long Short-Term Memory Neural Network (LSTM) or transformers generally expect a fixed number of time points from which to learn temporal relationships. Traditionally, to deal with the irregular time series nature of EHRs, many machine learning practitioners employ a method known as zero-padding. This involves appending zeros (or in some cases, prepending them) to the beginning and / or end of the time series data in order to create inputs of fixed size^21^. While zero padding is a standard technique for dealing with missing time series data, there are a variety of issues that can arise. The introduction of zeros can create artificial temporal relationships that do not actually exist.

Other methods average the patient notes over time, then combine it with a time-series embedding to obtain the final representation^15^. Additionally, machine learning architectures such as convolutional neural networks (CNN) and CNN-RNN architecture with semantic and temporal blocks can be leveraged to address temporality.^5,10^ More recently, transformers have been adapted for temporal data, such as the Flexible Time-aware LSTM Transformer, which incorporates a time aware mechanism.^14^ This mechanism addresses the irregular timing of notes by learning a flexible time decay function, effectively handling the variability in data spacing.

Large Language Models (LLMs) are artificial intelligence systems trained on extensive text data to understand and generate human-like language. These models have demonstrated remarkable capabilities in text generation, utilizing their comprehensive knowledge and understanding of language patterns. Recent works have leveraged LLMs for clinical applications, including named entity recognition, label generation, relation extraction, etc.^21,22^

We propose that LLMs, by harnessing knowledge from their extensive training data and the existing data in the EHR, will be capable of making meaningful observations that consider the temporal aspect of EHR data. LLMs have been found to be capable of providing information about a hypothetical datapoint given information about its embedding space^23^. Thus, by *filling in the gap* using text generated by LLMs, we hypothesize that we will be able to increase the accuracy of clinical predictions compared to existing machine learning and imputation methods for clinical text. Furthermore, through LLM generation, we can potentially mitigate disparities resulting from dataset irregularities and missing data, which can arise when certain groups have differing health status or limited access to healthcare. We leverage a variety of LLMs, including those trained specifically on biology and clinical data, to enhance the temporal structure of clinical notes from EHRs data. We feed the enhanced temporal structure into supervised deep learning models to predict 1-year mortality in ICU / ED patients. We compare to existing methods such as multimodal imputation, last observation carried forward (LOCF), and zero padding.

## Methods

### Data

We use the Medical Information Mart for Intensive Care IV (MIMIC-IV) database, which contains de-identified health data for patients who were admitted to either the emergency department or stayed in critical care units of the Beth Israel Deaconess Medical Center in Boston, Massachusetts^24^. MIMIC-IV excludes patients under 18 years of age. Date of death is derived from hospital and state records collected two years after the last patient discharge.

We restrict patients to those with at least two medical visits within 1-2 years apart. Visits occurring more than two years after the first visit are treated as a separate patient. A patient’s observation window is made up of medical visits that occur within two years after their first visit. We incorporate a 30-day buffer between the observation window and the prediction window so as not to use any visits where the patient may have died during (these patients were omitted). This process yields 33,123 patients with 12 percent of patients having experienced 1-year mortality. We then downsample randomly for a more balanced cohort (50/50). This yields 6070 patients (Table 1).

**Table 1:** Characteristics of our MIMIC-IV patient cohort.

|  |  | All | 1-year mortality + | 1 year mortality - |
| --- | --- | --- | --- | --- |
| All patients | Number of patients | 6070 | 3035 | 3035 |
|  | Average number of visits | 4.22 | 4.62 | 3.67 |
| Sex | Female | 3124 (0.52) | 1440 (0.47) | 1684 (0.55) |
|  | Male | 2946 (0.49) | 1595 (0.53) | 1351 (0.45) |
| Race | White | 4303 (0.71) | 2282 (0.75) | 2021 (0.67) |
|  | Black | 981 (0.16) | 422 (0.14) | 559 (0.18) |
|  | Hispanic | 307 (0.05) | 105 (0.03) | 202 (0.07) |
|  | Asian | 169 (0.03) | 88 (0.03) | 172 (0.03) |
|  | Other | 310 (0.06) | 138 (0.05) | 81 (0.06) |
| Age | 18-40 | 716 (0.12) | 105 (0.03) | 611 (0.20) |
|  | 41-65 | 2342 (0.39) | 977 (0.32) | 1365 (0.45) |
|  | 65-85 | 2335 (0.38) | 1444 (0.48) | 891 (0.29) |
|  | 85+ | 677 (0.11) | 509 (0.17) | 168 (0.06) |

From each patient visit we extract the first 512 words from each discharge summary (deleting chunks of text related to physical exam and discharge instructions). We use BioClinicalBERT to create a 768-dimensional vector representation of each discharge summary from each visit. BioClinicalBERT was initialized with BioBERT (trained on PubMed articles) and trained on all the notes from MIMIC-III, a past version of our database containing electronic health records from ICU patients from BIDMC between 2001 and 2012^9,25^.

### Zero padding

The use of zero-padding is a standard technique to deal with missing data in time series analysis, as it allows machine learning models to handle variable-length sequences while maintaining a consistent input size^26^. The maximum number of visits for a patient in our data is 79. Thus, for each patient, we fill in any missing doctor’s notes and ensure representation for up to 79 visits by using zero vectors of dimension 768 in accordance with the BioClinicalBERT embedding size (Figure 2a).

**Figure 2:**
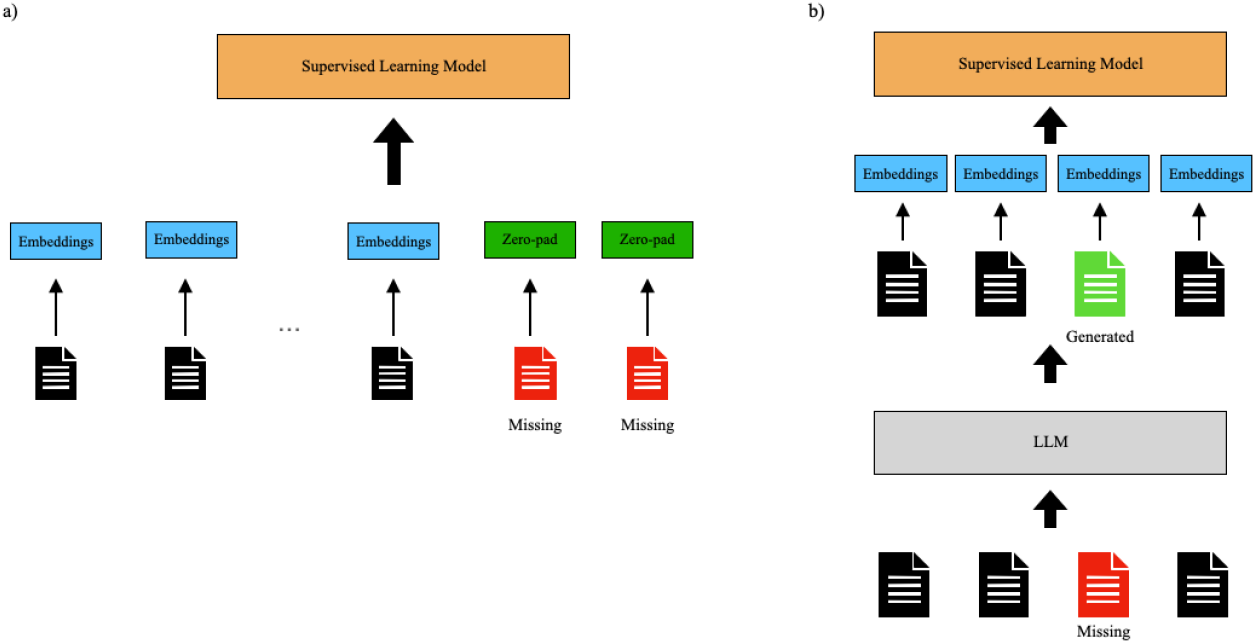
Illustration of zero-padding and LLM-based imputation: (a) BioClinicalBert embeddings are generated based on existing notes and zero-vectors are used for missing notes. (b) The LLM is given existing notes and asked to generate text for the missing notes. BioClinicalBert embeddings are calculated for both existing and generated text.

### Last Observation Carried Forward (LOCF)

For the rest of the temporal harmonization methods, we first apply temporal windows to the data. We consider the first visit as our index to align each patient, and split each patient record into four six-month intervals. If multiple visits occur in an interval, we keep the notes from the most recent visit. Naturally, if no visits occur in a particular interval, there is a missing gap. The following methods describe the different ways to fill that gap.

For each missing doctor’s note embedding, if it is not associated with the first visit, we reference the last observation for that patient and carry that embedding forward. In the case that the first visit is missing, we impute with a zero vector. This method is based on the rationale that the most recent observation for the patient typically holds the most relevant information about their condition.

### Multimodal Imputation

Supervised learning models may learn better by leveraging multimodal data to pull information from similar patients. MIMIC-IV data also provides codified data and structured data such as lab and vitals. Thus, we extract the diagnosis codes, prescription codes, procedure codes, and 30 most common labs and vitals in addition to the discharge summaries. We hypothesize that for two multimodal examples, if the majority of their modalities exhibit high similarities, then it is likely that their remaining modalities will also be similar. Thus, we propose that we leverage the most similar datapoint across the known modalities to impute the missing data in the remaining modality. Note that in this process, we mask out all note embeddings belonging to the same patient so that the algorithm is forced to carry over note embeddings from a different patient. We use cosine similarity for the similarity metric.

### Large Language Models

Finally, LLMs trained on large amounts of text data with billions of parameters may provide additional knowledge to help fill in the gaps between patient visits. We leverage LLMs to generate text data for the missing doctor’s notes based on the available doctor’s notes for that patient (Figure 2b). We compare the existing state-of-the-art LLM Generative Pre-trained Transformer 4 (GPT-4) to LLMs that were trained specifically on biology data and clinical data.

GPT-4 is a LLM developed by OpenAI, based on the transformer architecture. It demonstrates human-like language abilities and is trained on a large corpus of text data in an unsupervised manner^27^. BioMistral is an open-sourced LLM that uses Mistral as its foundation model and is further pre-trained on PubMed Central^28^. Asclepius is a specialized clinical large language model that is trained on synthetic clinical notes and further evaluated with real clinical notes from MIMIC-III discharge summaries. Asclepius has exhibited comparable performance metrics to GPT-3.5-turbo on several clinical benchmarks^29^.

To evaluate the performance of these three LLMs, we begin by exploring their capabilities in zero-shot learning. Zero-shot learning refers to the ability of a model to perform a task without having seen any explicit examples of that task during training^30^. We provide the LLM with the patient’s four visits, labeling any missing visits as ‘MISSING.’ We inform the LLM that each visit occurs at six-month intervals. We prompt the LLM to generate a doctor’s note for the missing visit about the patient’s symptoms and treatment plans based on the information from the existing visits.

We further explore few-shot learning, where the LLM is provided with a few examples of the task within the prompt itself, along with the original task instructions. This approach has been found to enhance the model’s performance by providing contextual examples^30^. However, few-shot learning may be limited by the LLM’s input size constraints, particularly when the original prompt is lengthy. Thus, we experimented with one-shot learning and modified the format of the example prompt. In this format, the example prompt includes notes from a single visit and the corresponding answer is the note from the subsequent visit occurring six months later. Following this, the original task instruction is provided.

### Supervised learning

We compare various methods for temporal harmonization of clinical text across multiple medical visits. For unstructured text data, we use BioClinicalBERT to extract embeddings. Due input size constraints of the BioClinicalBERT model, we truncate the text to the last 512 tokens and output a 768-dimensional vector representation of the text. All input text is lowercase.

In order to capture the temporal nature of the data, we use a BiLSTM model and a transformer model with positional encoding for evaluation. After the missing doctor’s notes have been filled through various methods, we extract the BioClinicalBERT embeddings then feed in the vectors into the models to predict mortality. The BiLSTM is trained for 100 epochs with an Adam optimizer and a learning rate of 1e-4 and consists of two layers with a hidden dimension of 256. The transformer architecture is also trained for 100 epochs and consists of one layer with four attention heads with an Adam optimizer and a learning rate of 1e-4.

### Algorithmic Fairness

We further assess our best method’s performance by segmenting the data based on the percentage of missing information. We compare the effectiveness of zero-padding, and the best temporal harmonization methods across varying levels of data incompleteness.

## Results

We evaluate the performance of various temporal harmonization methods by comparing their area under the curve (AUC) scores and F1 metrics for both BiLSTM and transformer architectures. In both architectures, GPT-4 with zero-shot prompting achieves the highest AUC score. For F1-score, GPT-4 with zero-shot prompting leads in the BiLSTM architecture, while GPT-4 with one-shot prompting has the best performance with the Transformer architecture. GPT-4 significantly surpasses the baseline zero-padding method (Table 2, Figure 3). For precision and recall, see Table S1 in the supplement.

**Table 2:** Performance metrics of supervised learning methods by temporal harmonization method.

|  | BiLSTM |  | Transformer |  |
| --- | --- | --- | --- | --- |
|  | AUC | F1 score | AUC | F1 score |
| **Zero pad** | 0.739 | 0.715 | 0.797 | 0.460 |
| **LOCF** | 0.748 | 0.679 | 0.791 | 0.649 |
| **Multimodal Imputation** | 0.683 | 0.642 | 0.725 | 0.448 |
| **Biomistral (zero shot)** | 0.739 | 0.690 | 0.770 | 0.571 |
| **Biomistral (one shot)** | 0.733 | 0.674 | 0.777 | 0.715 |
| **Asclepius (zero shot)** | 0.741 | 0.690 | 0.769 | 0.640 |
| **Asclepius (one shot)** | 0.742 | 0.669 | 0.774 | 0.726 |
| **GPT-4 (zero shot)** | **0.785** | **0.723** | **0.817** | 0.706 |
| **GPT-4 (one shot)** | 0.777 | 0.706 | **0.817** | **0.734** |

**Figure 3:**
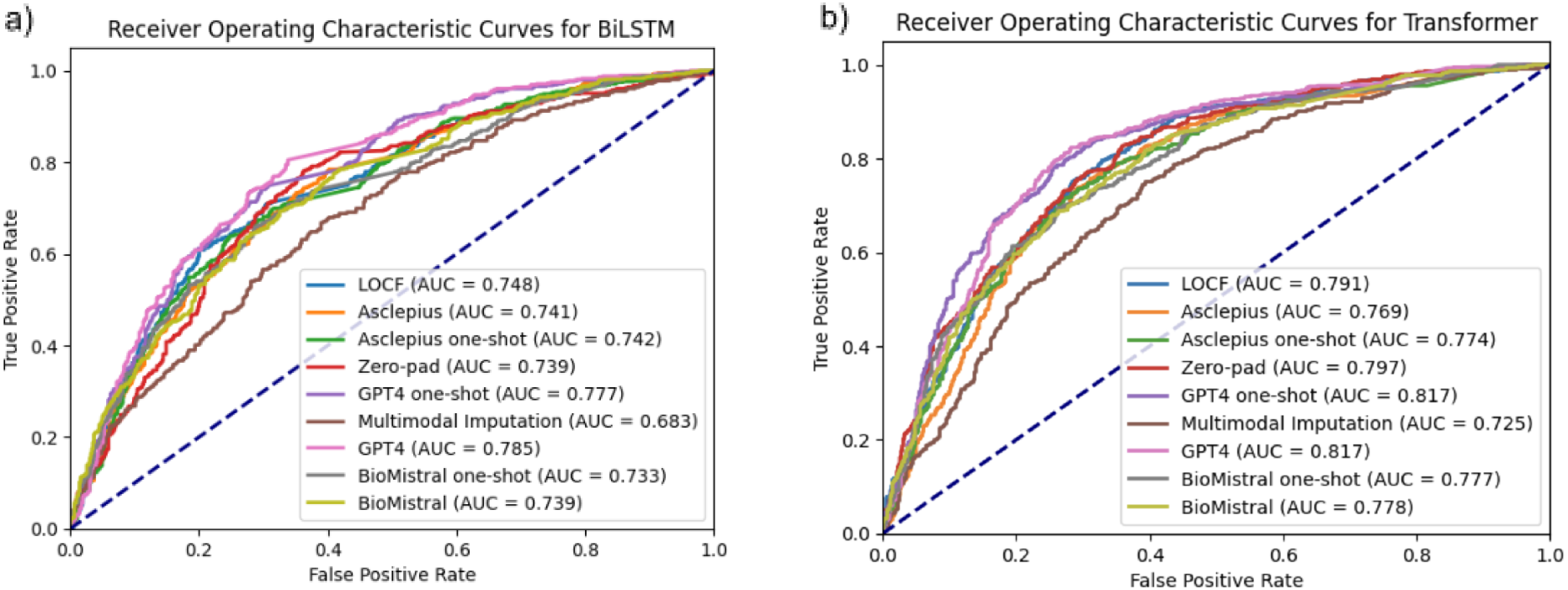
ROC curves by temporal harmonization method for a) BiLSTM and b) Transformer

We find that there is no significant difference in performance between zero-shot prompting and one-shot prompting across all three LLMs. When we evaluate the AUC scores and F1-metric, there is no clear trend that suggests that one-shot prompting increases the accuracy of the generated text (Table 2, Figure 3).

For non-LLM methods, we see that the LOCF approach shows a slight improvement over the baseline method of zero-padding. We further note that Multimodal Imputation underperforms compared to the baseline method of zero-padding (Table 2, Figure 3).

After applying temporal windows on the data, each patient has four time intervals that occur six months apart. After identifying the best performing temporal harmonization methods, we stratify our test data by patients with less than 50% missingness for those time intervals, and those with more than 50%. We observe huge improvements in model performance for both BiLSTM (AUC from 0.728 to 0.846, F1 score from 0.673 to 0.717) and transformer (AUC from 0.781 to 0.836, F1 score from 0.608 to 0.705) when using GPT-4 zero shot and one shot prompting to fill in the gaps (Table 3). For precision / recall results, see Table S2 in the Supplement.

**Table 3:** Quantifying improvements in performance for patients with more or less missing data.

|  |  | BiLSTM |  | Transformer |  |
| --- | --- | --- | --- | --- | --- |
|  |  | AUC | F1 score | AUC | F1 score |
| **Missing < 50%** | **Zero pad** | 0.749 | 0.712 | 0.801 | 0.467 |
|  | **GPT-4 (zero shot)** | 0.809 | 0.747 | 0.808 | 0.744 |
|  | **GPT-4 (one shot)** | 0.761 | 0.711 | 0.801 | 0.724 |
| **Missing > 50%** | **Zero pad** | 0.728 | 0.673 | 0.781 | 0.608 |
|  | **GPT-4 (zero shot)** | 0.846 | 0.739 | 0.818 | 0.717 |
|  | **GPT-4 (one shot)** | 0.813 | 0.717 | 0.836 | 0.705 |

## Discussion

In this work, we demonstrated the effectiveness of GPT-4 for temporal harmonization in supervised learning models when modeling medical notes for clinical prediction. Since GPT-4 is not an open source model, it is unknown what data were used to train it. However, its performance in temporal harmonization indicates that it was likely trained on medical data along with other sources of data to provide insightful predictions about patient conditions when filling in the missing gaps. Particularly for patients with less medical visits, we observe huge improvements in model performance by filling those gaps with text generated by GPT-4. Previous studies have shown that missing data in EHRs can negatively impact model performance in disease prediction models^19^. In the emergency department / ICU case, patients with less visits are more likely to be healthier. However, in a primary care setting, underserved populations are more likely to have less data in their record. These results display the potential for GPT-4 to combat algorithmic unfairness by augmenting the EHRs for patients with less data.

Interestingly, our results indicate that the use of GPT-4 for temporal harmonization in clinical prediction models surpasses the use of clinical and biological LLMs in performance, highlighting the adaptability of GPT-4 for healthcare applications. This suggests that even without targeted training in medical text, GPT-4 is able to generalize and provide relevant information in various clinical scenarios. This suggests exciting new directions for research in applying more generalist LLMs to healthcare, and identifying different scenarios where one may or may not need specific clinical LLMs.

We observe that Multimodal Imputation performs worse in our supervised deep learning models than the baseline method of zero-padding. This is likely due to the fact that Multimodal Imputation masks out note embeddings from the same patient during imputation. Even if data from the other modalities is very similar, forcing the imputation to carry over note embeddings from a different patient could introduce noise into the note embeddings.

We further note that one-shot prompting did not lead to a consistent increase in model performance compared to zero-shot prompting. This may be an indicator that the LLM models have strong generalization capabilities even without task-specific training. In that case, we could come to the conclusion that zero-shot prompting is sufficient for our setting. However, the lack of performance improvement could also be attributed to the length of the prompts and the potential limitations of the LLM in handling longer prompts. Additionally, the discrepancy in formatting between the example prompt and the actual prompt, motivated by constraints on input length, may have diminished the impact of one-shot prompting.

There are a few limitations to this study. Future research could focus on enhancing explainability with LLMs and reducing instances of hallucinations. Currently, due to the black-box nature of LLMs, it is difficult to determine how they arrive at specific outputs, which is problematic in the healthcare sector where transparency is critical. We further anticipate that improving explainability in LLMs could also contribute to minimizing hallucinations in their outputs. Therefore, research into methods of identifying and correcting hallucinations in LLM outputs will be a significant step towards their safe and effective use in the domain of healthcare. Finally, it would be worth investigating the impact of LLMs on temporal harmonization of primary care data, or even *simulating* missingness in ICU clinical notes that reflect barriers in access to healthcare (similar to Getzen et al., 2023), and evaluating LLMs’ abilities to fill in the gaps and improve clinical prediction performance.

## Data Availability

All data produced are available online at https://physionet.org/content/mimiciv/2.2/

## Acknowledgements

This work was supported in part by National Institutes of Health grants, U01CA274576 and RF1AG063481. The content is solely the responsibility of the authors and does not necessarily represent the official views of the National Institutes of Health.

## Supplementary material

**Table S1:** Performance metrics of supervised learning methods by temporal harmonization method including PPV and Recall

|  | BiLSTM |  |  |  | Transformer |  |  |  |
| --- | --- | --- | --- | --- | --- | --- | --- | --- |
|  | AUC | PPV | Recall | F1 score | AUC | PPV | Recall | F1 score |
| **Zero pad** | 0.739 | 0.671 | **0.764** | 0.715 | 0.797 | **0.824** | 0.318 | 0.460 |
| **LOCF** | 0.748 | 0.689 | 0.671 | 0.679 | 0.791 | 0.756 | 0.568 | 0.649 |
| **Multimodal Imputation** | 0.683 | 0.617 | 0.669 | 0.642 | 0.725 | 0.696 | 0.331 | 0.448 |
| **Biomistral (zero shot)** | 0.739 | 0.645 | 0.742 | 0.690 | 0.770 | 0.776 | 0.452 | 0.571 |
| **Biomistral (one shot)** | 0.733 | 0.671 | 0.678 | 0.674 | 0.777 | 0.651 | 0.793 | 0.715 |
| **Asclepius (zero shot)** | 0.741 | 0.663 | 0.720 | 0.690 | 0.769 | 0.737 | 0.566 | 0.640 |
| **Asclepius (one shot)** | 0.742 | 0.695 | 0.646 | 0.669 | 0.774 | 0.682 | **0.776** | 0.726 |
| **GPT-4 (zero shot)** | **0.785** | **0.713** | 0.736 | **0.723** | **0.817** | 0.790 | 0.638 | 0.706 |
| **GPT-4 (one shot)** | 0.777 | 0.709 | 0.703 | 0.706 | **0.817** | 0.767 | 0.703 | **0.734** |

**Table S2:** Quantifying improvements in performance for patients with more or less missing data including PPV and Recall

|  |  | **BiLSTM** |  |  |  | **Transformer** |  |  |  |
| --- | --- | --- | --- | --- | --- | --- | --- | --- | --- |
|  |  | AUC | PPV | Recall | F1 score | AUC | PPV | Recall | F1 score |
| **Missing < 50%** | **Zero pad** | 0.749 | 0.727 | 0.697 | 0.712 | 0.801 | 0.826 | 0.326 | 0.467 |
|  | **GPT-4 (zero shot)** | 0.809 | 0.747 | 0.747 | 0.747 | 0.808 | 0.728 | 0.759 | 0.744 |
|  | **GPT-4 (one shot)** | 0.761 | 0.715 | 0.705 | 0.711 | 0.801 | 0.771 | 0.683 | 0.724 |
| **Missing > 50%** | **Zero pad** | 0.728 | 0.670 | 0.676 | 0.673 | 0.781 | 0.753 | 0.509 | 0.608 |
|  | **GPT-4 (zero shot)** | 0.846 | 0.757 | 0.722 | 0.739 | 0.818 | 0.731 | 0.704 | 0.717 |
|  | **GPT-4 (one shot)** | 0.813 | 0.681 | 0.750 | 0.717 | 0.836 | 0.800 | 0.629 | 0.705 |

